# A Pre-Post Design Exploring the Potential Benefits of a Balance Enhancement Program for Deaf Older Adults: A Pilot Study

**DOI:** 10.64898/2026.11.17.26368347

**Authors:** Carol Cutler Riddick, Chizuko Tamaki

## Abstract

**Background:** Deaf older adults are at a heightened risk for falling, yet no fall-reduction programs for Deaf persons have been reported. Culturally responsive balance enhancement programs targeting this underserved population are needed. Consequently, the purposes of the study were to develop and pilot an exercise program for older adults who are Deaf and use American Sign Language (ASL).

**Methods:** During a two-year period, 40 community-dwelling Deaf adults (25 women, 15 men; ages 62-92 years) attended two 2-hour classes each week for eight weeks on a college campus, delivered entirely in ASL, with an additional expectation to engage in exercises outside of class. Outcome measures completed at pre– and post-intervention were timed up-and-go (TUG), sit-to-stand (STS), chair sit and reach (SR), Activities-specific Balance Confidence (ABC), and Fall Risk Questionnaire (FRQ). Theoretical frameworks shaped the content and delivery of BIM.

**Results:** 67.5% of participants attended ≥14 classes. Paired-samples t-tests revealed significant improvements in TUG, STS, and SR. Participants who entered the program with an ABC score below 90% experienced substantial improvement, while score changes on the FRQ were not statistically significant even among the at-risk group. Eight adverse events (seven falls) occurred in class, with no serious consequences.

**Conclusion:** The culturally responsive BIM engaged participants effectively and led to significant improvements across multiple measures of dynamic balance. Given the high rates of vestibular and lower body weakness among Deaf older adults, vigilant management of fall risks is needed for future class offerings.

## Background

For persons in the sixth decade of life and older, falls can be a life-changing, if not a life-ending event. One in three older adults fall each year (Dolan et al., 2025), resulting in falls being the leading cause of injury and death for persons age 65 years and older (Garnett et al., 2025). Reducing falls and fall-related injuries and deaths of older adults has become a recognized public health priority, as evidenced by the injury prevention initiatives of the Centers for Disease Control and Prevention (CDC, 2026), the national health objectives established in Healthy People 2030 (U.S. Department of Health and Human Services [HHS], n.d.), and the World Health Organization’s (WHO) designation of falls as a major global public health problem (WHO, 2021).

Systematic analyses have identified non-modifiable and modifiable fall risk factors that exist for community-dwelling older adults (e.g., Deandrea et al., 2010; Pillay et al., 2024). Non-modifiable fall risk factors that have emerged include advancing age, history of falls, and underlying sensory or neurodegenerative declines (e.g., hearing loss, vestibular loss, vision loss, cognitive impairment, Parkinson disease). Contrastingly, efforts to improve modifiable physiological and physical performance factors, such as gait mechanics, functional ability, and lessening extremity muscle weakness, have been linked to improved balance. Further mutable contributors to falls include medications that cause dizziness, environmental factors (such as a slippery surface or cluttered floor), urgency or rushing (e.g., to the bathroom), and fear of falling.

Numerous interventions have been studied to ameliorate fall risks, such as education, exercise, medication review, home hazard assessment, and psychological interventions. A meta-analysis of 21 interventions reported that 14 with the most promise had a focus on exercise (Pillay et al., 2024). For these clusters of studies, exercise interventions were supervised and fall incidence was monitored for at least three months. Other exercise efficacy reports note exercise interventions result in improvements on an array of measures, including flexibility, coordination, lower limb strength, balance, and decreased fear of falling (Adeyemi et al., 2025; Deck et al., 2026; Sadaqa et al., 2023). Moreover, exercise interventions to improve balance and reduce fall risk, particularly in group settings, have revealed additional benefits, such as the enjoyment of social or peer support (Hernán-Santana & Sobrido-Prieto, 2026; Peng et al., 2026).

The odds of falling are up to 50% higher among people with hearing loss (Lin & Ferrucci, 2012; Yeo et al., 2025), yet the efficacy of balance enhancement or fall reduction interventions for culturally Deaf persons, who typically present with severe and profound ranges of hearing loss, has not been studied. A few programs have reported the intentional sampling of older adults with audiological hearing loss, but those targeting culturally Deaf older adults are non-existent (Jones et al., 2019; Lambert et al., 2017; Wollesen et al., 2021).

Identifying oneself as *Deaf,* with the use of capital “D,” connotes that a person is affiliated with the community that embraces their cultural heritage. While Deaf communities are diverse, including in terms of their audiologic presentations, American Sign Language (ASL) is the cultural thread that binds Deaf communities in the United States. Instead of viewing being audiological-defined deaf or hard-of-hearing from a biomedical disability-based perspective (cf. Koon et al., 2025), being Deaf is seen from a socio-cultural viewpoint (Krawczyk et al., 2024).

Addressing Deaf older adults’ needs in a culturally responsive manner is particularly important in improving this population’s health outcomes. From a communication standpoint, when health-related services are delivered by a provider who uses the same language as that of the patients’, improved health outcomes have become evident a majority of the time (Diamond et al., 2019). While many Deaf individuals are proficient in written or oral English, communicating in ASL enhances cognitive processing abilities, enables authentic self-expression, promotes social connectedness, builds trust through cultural belonging, and fosters peer support (McKee et al., 2012). In a group setting intervention, building communication and trust extends beyond provider-participant relationships; peer support and social engagement benefits are severely limited if participants cannot communicate with each other without barriers.

Aside from language, there are Deaf community-specific communication norms that ensure visual access to information (Bauman, 2014). These include physical environment considerations such as furniture and equipment setup that does not limit lines of sight to all activities held in the space, adequate lighting that is not harsh or glaring, seating arrangements that allow all participants to see each other; conversation turn-taking practices that allow participants time to locate and visualize the speaker who holds the floor; and sequential rather than simultaneous instructions where instructions are always provided in full before the participants’ visual attention moves away from the instructor.

Physiologically, the high rate of severe vestibular impairment among Deaf older adults (Tamaki & Cole, 2026) makes them highly somatosensory dependent during exercises. When individuals have severe vestibular limitations, quick head movements do not generate automatic stabilizing motor responses. Consequently, individuals must move at a speed at which proprioceptive and visual feedback can be processed and used for stabilization. Ample time is needed to make transitions between instructions and exercise moves so that the head velocity can remain low. Somatosensory inputs must be ensured through maintaining an adequate base of support, with supportive equipment such as bars and stable chairs nearby.

Clearly, any intervention that attempts to improve balance must be designed in a manner responsive to the vestibular dynamics and cultural mores of older Deaf individuals. Thus, the efficacy of these tailor-made interventions must be examined and reported. Indeed, Freiberger (2026) has implored investigators to disseminate their innovative fall research.

### Objectives

The overarching purposes of the project were two-fold. First was to create a series of written lesson plans for a balance-enhancement exercise program that is designed to specifically address the needs of ASL-using Deaf older adults. The *Balance in Motion (BIM) program* was designed to be practiced both in group and home settings (Montero-Odasso et al., 2022). Second was to pilot BIM with Deaf people to determine feasibility, effectiveness, and further refinement needs.

## Methods

### Balance in Motion Curriculum Development

Four theoretical frameworks steered the project. Namely, BIM’s development, design, and implementation were linked to the *Social Ecological Model* (McLeroy et al., 1988), the United Nation’s *Healthy Ageing Plan of Action 2020-2030* (WHO, 2020; 2023), the components of exercise training recommendations by the American College of Sports Medicine (2021), and *Teaching-Rules-Environment-Equipment Inclusiveness Paradigm* (Australian Sports Commission, n.d.).

The Social Ecological Model provides a framework to describe how BIM evolved in order to improve the balance of Deaf older adults (McLeroy et al., 1988: Sallis et al., 2015). This framework provided insights into the interpersonal, cultural, linguistic, cognitive, and environmental factors that should be considered when designing an exercise and fall risk reduction program for Deaf older adults. At the organizational level, faculty within a university community serving Deaf and Hard of Hearing students coalesced to design a balance-enhancing intervention, drawing on the institution’s mission and its existing relationships with the population it serves. Content and delivery were driven by translating, applying, and testing state-of-the-art knowledge discoveries into an intervention that was mindful and responsive to Deaf older adults. The Social Ecological Model guided the development and implementation of the BIM program in terms of creating an environment that delivered content, in a linguistically appropriate manner, that fostered social support and promoted balance confidence among the participants.

The World Health Organization’s (2021) *Decade of Healthy Aging* proclamation also contributed to the development of BIM. Among other things, this pronouncement calls for delivering responsive, person-centered services. One service mechanism singled out to promote healthy aging is the creation of physical activity programs that promote functional abilities and social interactions during physical exercise and recreation activities.

The organization of BIM workouts was developed according to *Exercise Training Component Principles* recommended by the American College of Sports Medicine (2021, p. 153). Each exercise session had three phases (each meeting prescriptive standards for number of sets and repetitions): Dynamic warm-up stretching (5-10 minutes), conditioning exercises (30 minutes), and static cool-down stretching (5-10 minutes).

BIM’s content and delivery were also informed by the *Teaching-Rules-Environment-Equipment (TREE) Inclusiveness Philosophy* (Australian Sports Commission, n.d.). Thus, BIM recognized the varying abilities of participants by providing adaptations in Teaching Style (e.g., bottom-up skill development, proprioceptive drills), Rules (e.g., decreased ball/birdie boundaries, number of bounces allowed), Equipment (e.g., soft grip rackets, larger balls/birdies), and Environment (e.g., minimized lighting glare, unobstructive eyesight lines). Two Deaf and one hard-of-hearing faculty members in the Sports, Recreation, and Fitness Program, each of whom with unique specialized credentials related to exercise facilitation, contributed to the creation of the lesson plans for exercises and adapted recreation activities.

The substance of the lessons was pulled primarily from Harvard Health Publishing’s (HHP) *Special Health Reports* on better balance and fall prevention (HHP, 2022; 2024b; 2024c), stretching (HHP, 2023), strength and power training (2019), walking (2015), and functional fitness (2024a). Special attention was paid to ensure each exercise was functional (Liu et al., 2024). Movements were selected with the purpose of improving the ability to safely perform both activities of daily living (e.g., using the toilet, bathing) and carrying out instrumental activities needed for independent living (e.g., reaching a higher shelf or bending down lower to retrieve objects; lateral moves and changing directions such as in a kitchen or bathroom). A sampling of the thematic topics used for exercise classes were Walking, Fitness Walking; Chair & Standing Pilates; Lower Body Strength; Steps, Curbs & Trip Recovery; Getting Up & Down from Chair and Floor, and Moves Can Do Anywhere: Driving, Standing in Line, Cooking.

### Pilot of Balance in Motion Program

#### Design

A single group pre-post intervention design was used. Participants were enrolled in one of the four eight-week BIM programs offered between the fall 2024 and spring 2026 semesters, held in person at the Gallaudet University campus in Washington, DC.

The study was approved by the author institution’s Institutional Review Board for Human Subject Research (IRB-FY24-34). The study was registered, and its protocol can be accessed through ClinicalTrials.gov (NCT06912607) prior to participant enrollment.

#### Participants

Potential participants were recruited by various means, including fliers, videos, and announcement blurbs distributed on campus, through alumni networks, and regional Deaf organizations. Additionally, recruitment efforts extended to in-person outreach at Deaf organizations’ meetings as well as at campus homecoming and other on-campus athletic events. Participants also contributed to recruitment via word-of-mouth referral, leading additional individuals to volunteer for participation. For those expressing interest in becoming a study participant, a one-on-one meeting in the participant’s preferred mode (videophone, Zoom, in person) was scheduled to determine participation eligibility.

Eligibility was determined through an intake screening interview, conducted in ASL. Inclusion criteria were: (1) Self-identified as Deaf, (2) Used ASL, (3) Resided in a home, not in an institution, (4) Was able to walk 20 feet unassisted, and (5) Expressed no hesitation to attend at least 14 out of 16 sessions of BIM classes held on campus. Exclusion criteria were being blind or having been diagnosed with dementia. Additionally, while not an exclusion criterion, those with medical conditions that may impede exercise routines (i.e., cardiovascular, metabolic, or renal disease) were required to seek medical clearance from their healthcare provider prior to enrollment.

A target sample size of 40 was set based on available staffing to ensure participant safety. A priori power analysis conducted for paired t-tests using G*Power 3.1 (Faul et al., 2007, 2009) indicated that a sample size of 34 participants would be required to achieve a target statistical power (1-β) of 0.80, assuming a medium effect size (*d* = 0.50). For a power of 0.90, 44 participants would be needed.

#### Intervention Delivery

The BIM program was delivered using a three-part framework consisting of formal onsite exercise classes, adapted recreation activities, and home-based practice. The formal program spanned eight consecutive weeks with two 2-hour sessions per week (16 class meetings total). Each session dedicated the first hour to structured exercise and the second hour to an adapted recreation activity, including four sessions each of seated volleyball, ping pong, badminton, and pickleball, designed to promote dynamic balance through play while fostering social interaction and peer support.

Classes were conducted entirely in ASL. Each exercise session was co-taught by two or three faculty instructors who demonstrated movements alongside visual aids projected on a 72-inch monitor. Four to six additional team members were positioned throughout the room to reinforce proper technique, offer encouragement, and provide immediate balance support (e.g., standing in a ready position or placing a chair adjacent to participants). Undergraduate student interns led the adaptive recreation component under faculty supervision, maintaining the same safety protocols and staff-to-participant ratio.

To meet national physical activity guidelines (Bull et al., 2020; HHS, 2018) and encourage sustained habits after the program concluded, participants were expected to complete at least 50 minutes of self-directed practice at home each week using exercises learned in class. To facilitate home practice, participants received a laminated exercise card deck after each session. Each card featured color-coded anatomical diagrams illustrating targeted muscle groups and joints, step-by-step exercise routines with suggested adaptations, and a QR code linking to instructional videos of a Deaf team member demonstrating the movements in ASL.

#### Vestibular Functioning Preceding Program Participation

Vestibular function was assessed before the first BIM class to identify participants at risk of falling during class. The following measures were employed: (1) video head impulse test (vHIT; using Interacoustics EyeSeeCam), to assess the vestibulo-ocular reflex mediated by the lateral semicircular canals and the superior branch of the vestibular nerve; (2) cervical vestibular evoked myogenic potentials (cVEMP; using Intelligent Hearing Systems Aria), to evaluate the vestibulo-collic reflex mediated by the saccule (gravity-sensing vestibular organ) and the inferior branch of the vestibular nerve); and (3) the modified Clinical Test of Sensory Integration on Balance eyes-closed on foam condition (mCTSIB EC-F; using Bertec Essential Portable), to measure the sway velocity during a bilateral stance that forced a reliance on vestibular inputs due to restricted access to somatosensory and visual information. A participant was categorized as having a vestibular impairment if vHIT showed abnormal vestibulo-ocular reflex gains, cVEMP was absent in either ear, or the participant could not remain standing without stepping, opening eyes, or needing support for any of the three 10-second trials in mCTSIB EC-F. Participants with a vestibular impairment and/or severely limited lower body strength were subsequently assigned a staff member(s) to stand within arm’s reach of the participant during BIM classes, in order to provide a steady hand or bracing assistance as needed.

#### Outcome Variables

Five outcome variables were hypothesized to improve through the BIM program participation, and were administered at both pre– and post-intervention assessments: (1) Timed-up-and-go (TUG), an assessment of lower body strength, gait speed, and dynamic balance; (2) Sit-to-stand (STS), a test of lower body strength and endurance; (3) Chair sit and reach (SR), for lower extremity flexibility (extensibility of hamstrings and lumbar spine mobility); (4) Activity-specific Balance Confidence scale (ABC), a self-reported confidence in performing 16 activities; and (5) Fall-Risk Questionnaire (FRQ), a self-reported account of fall risk factors.

TUG was administered as typically described in the literature (e.g., CDC 2017b). The participant was instructed to rise from an armless chair without using hands to push off, walk along a 10-foot path, turn around at the 1-ft tall orange cone, walk back the 10-foot path, then sit down into the chair. A manual stopwatch was used to count the time elapsed to execute the task to the nearest second. STS was also administered as typically described in the literature (e.g., CDC 2017a), using an armless chair set up with its back placed against a wall for stability. With the arms folded in front, the number of full stand-and-sit repetitions completed within a 30-second period was recorded. The SR (Rikli & Jones, 2013) was administered with the participant sitting on the edge of a chair, extending one leg forward with the heel on the floor, and reaching both hands toward the toes. The distance between the toes and the tip of the fingers was measured in centimeters (cm). The validated ASL versions of ABC (Tamaki & Cole, 2026) and FRQ (Conrad et al., 2026) were administered through Qualtrics (Provo, UT).

Pre-intervention assessments were completed 1-2 weeks prior to the BIM’s first class, with one participant completing it 2 days before their first class. Post-intervention assessments were completed on the day of the last class meeting for 32 of the 40 participants. The remaining eight returned on a later date: six participants within a week, and two participants at 12 and 19 days.

#### Data Analysis

Data were analyzed for the 40 participants who completed the intervention. Descriptive statistics summarized demographic and vestibular status. Paired-samples *t*-tests were used to detect improvements from pre– to post-intervention measurements. Statistical significance was set at the *α* = .05 (or *p* < .05) one-tailed probability level. Statistical analyses were performed using Jamovi Version 2.7 (2024) and R version 4.5.1 (R Core Team, 2025).

## Results

### Characteristics of the Sample

Of the 48 volunteers who initially expressed interest, 40 completed the eight-week intervention. Seven volunteers did not begin the intervention. One additional volunteer began the intervention but did not complete it due to a medical condition unrelated to study participation. Figure 1 displays the participant tracking through recruitment, screening, enrollment, and completion.

**Figure 1.**
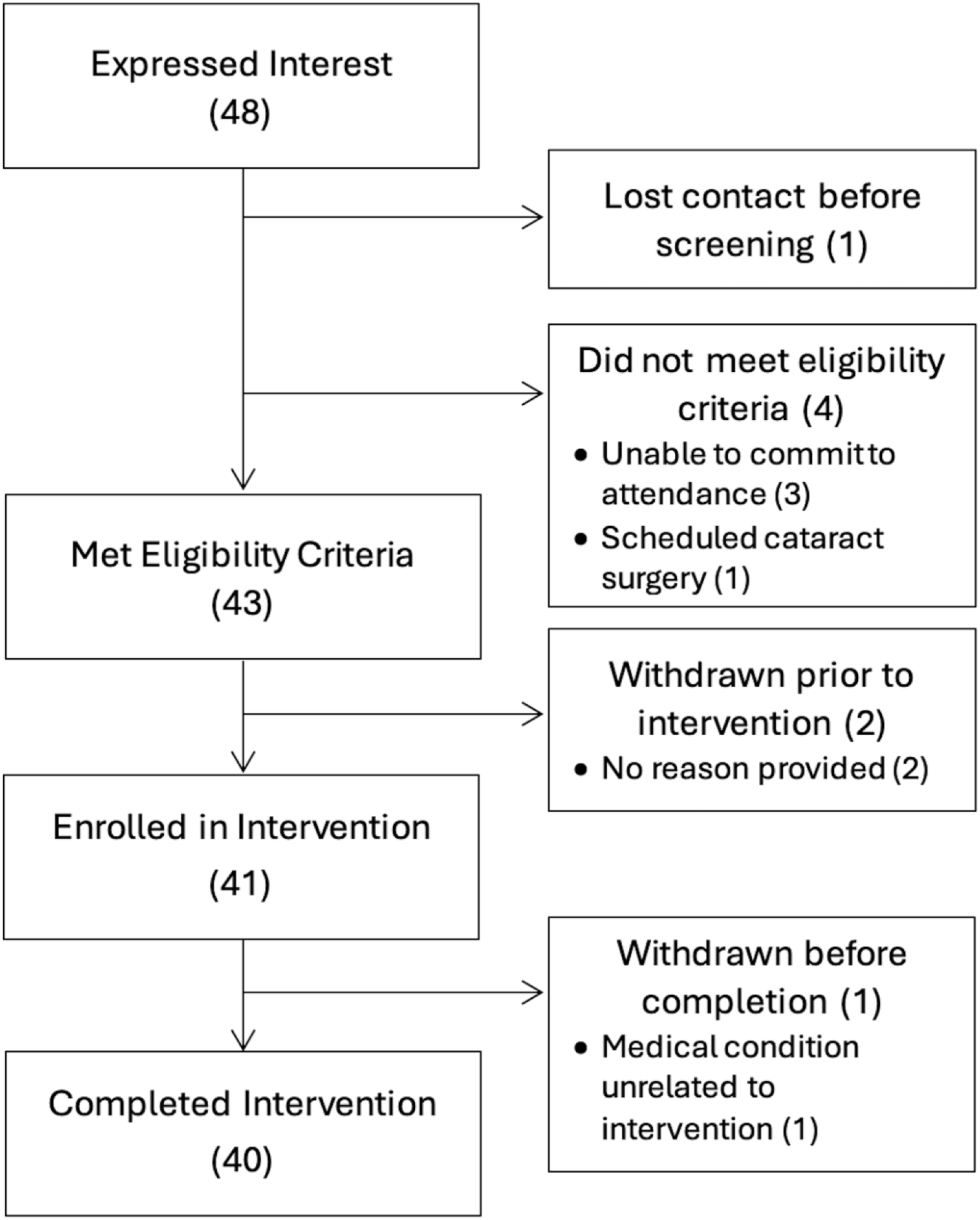
A flow diagram of the number of volunteers at recruitment, enrollment, and completion of the study.

Of the 40 participants who completed the intervention, 25 (63%) were women and 15 (37%) were men; 33 (72%) identified as being non-Hispanic White and 7 (28%) identified as Black, Asian, Native American, and/or Hispanic. The median age of study participants was 71 years (range 62-92 years). Pre-testing of vestibular function and lower body strength revealed significant physical limitations in the sample. Of 40 participants, 34 (86%) had vestibular function limitations, and 23 (58%) had lower body strength below age– and sex-adjusted norms (CDC 2017a). Nearly half of the participants (i.e., 19, or 48%) had a combination of both vestibular and lower body strength limitations. Table 1 summarizes the sample characteristics.

**Table 1.** Demographic and Pretest Characteristics of BIM Participants.

| **Cohort**<br>**(n)** | **Gender** |  | **Median**<br>**Age years**<br>**(range)** | **Pretest Condition/Limitations** |  |  |
|---|---|---|---|---|---|---|
|  | **Female**<br>**n (%)** | **Male**<br>**n (%)** |  | **Vestibular**<br>**Impairment**<br>**n (%)** | **Lower Body**<br>**Weakness**<br>**n (%)** | **Both**<br>**Limitations**<br>**n (%)** |
| Fall 2024<br>(n=6) | 5 (83%) | 1 (17%) | 79.5 (62-92) | 5 (83%) | 4 (50%) | 2 (33%) |
| Spring 2025<br>(n=12) | 4 (33%) | 9 (67%) | 68 (62-86) | 7 (58%) | 5 (42%) | 3 (2c5%) |
| Fall 2025<br>(n=12) | 8 (67%) | 4 (33%) | 70 (63-81%) | 12 (100%) | 7 (58%) | 7 (58%) |
| Spring 2026<br>(n=10) | 8 (80%) | 2 (20%) | 75.5 (65-86) | 10 (100%) | 7 (70%) | 7 (70%) |
| **TOTAL** | 25 (63%) | 15 (37%) | 71 (62-92) | 34 (86%) | 23 (58%) | 19 (48%) |

### Attendance, Compliance, and Adverse Events

The majority (27/40; 67.5%) of participants attended at least 14 of the 16 BIM sessions offered. Reasons provided for absences included inclement weather (ice, snow, storm forecast), broken vehicle/unavailable transportation; illness; scheduling conflicts with medical appointments; or prior travel commitments (visit family or vacation).

Participants often reported difficulty making or finding time to engage weekly in aerobic activity or practicing BIM content outside of formal class meetings. A variety of reasons were cited for non-compliance, including lack of a workout buddy, not adept at being able to access recorded summaries of *BIM* lessons using the provided QR codes, not having a safe place to walk outside, inclement weather, and illness.

There were eight in-class adverse event incidents: seven were falls and trips, and one was a ping pong ball hit to the face around an eye. All incidents occurred during the recreational activities, with the most frequent instances of falls happening when participants attempted to return a ball or birdie during ping pong, badminton, and pickleball. In seated recreation (i.e., badminton, pickleball), falls ensued when participants either stood up in excitement or when they remained seated while attempting to reach a shuttle beyond their reach. There was one instance of a fall due to a participant tripping on badminton netting equipment. None of these incidents resulted in a serious injury, meaning the participant reported no injury and returned to the activity within a few minutes, or had a minor injury that, based on self-report, required some rest and icing for up to three days. Participants were followed up for a week to confirm that there were no lasting or delayed-onset problems.

Aside from these specific fall incidents, multiple participants reported episodes of muscle soreness the day following a BIM class meeting. Additionally, one person, due to chronic fatigue, spent about half of the time sitting during the adaptive recreation activity period.

### Outcome Variables

Both baseline and post-intervention measures of TUG, STS, and SR were available from all 40 participants. Baseline ABC and FRQ were missing from one participant; thus, these analyses were performed based on 39 participants.

Paired samples *t*-test indicated significant improvements with large effect sizes in TUG (*t* = – 5.91, *p* < .001, *d* = 0.93) and STS (*t* = 4.83, *p* < .001, *d* = 0.76), and a significant improvement with a small effect size for SR (*t* = 2.05, *p* = .024, *d* = 0.32). Contrastingly, *t*-test did not detect significant changes in participants’ self-reports on balance confidence (ABC, *t* = 0.92, *p* = .181, *d* = 0.15) or risk of falling (FRQ, *t* = –0.52, *p* = .304, *d* = 0.08). Table 2 presents the results.

**Table 2.** Paired Samples T-Test Comparing Pre– and Post-Assessments.

| **Test** | ***M<sub>Pre</sub>* (SD)** | ***M<sub>Post</sub>* (SD)** | **SE<sub>M diff</sub>** | ***t* (df)** | ***p* (1-tail)** | **Effect size (Cohen’s *d*, 95% CI)** |
|---|---|---|---|---|---|---|
| **TUG** (s) | 10.9 (3.63) | 8.3 (3.86) | 0.440 | -5.91 (39) | <.001* | 0.93 (0.56-1.20) |
| **STS** (reps) | 9.7 (4.04) | 12.1 (5.15) | 0.486 | 4.83 (39) | <.001* | 0.76 (0.41-1.1) |
| **SR** (cm) | -4.8 (11.63) | -1.0 (9.18) | 1.855 | 2.05 (39) | .024* | 0.32 (0.0-0.64) |
| **ABC** (%) | 85.2 (18.66) | 86.5 (17.15) | 1.375 | 0.92 (38) | .181 | 0.15 (-0.17-0.46) |
| Baseline <90% | 73.2 (22.0) | 79.4 (22.60) | 1.900 | 3.30 (17) | .002* | 0.78 (0.24-1.30) |
| **FRQ** (points) | 4.1 (3.19) | 3.9 (3.23) | 0.346 | -0.52 (38) | .304 | 0.08 (-0.40-0.23) |
| Baseline ≥4 points | 6.5 (2.52) | 5.6 (3.30) | 0.523 | -1.72 (19) | .051 | 0.39 (-0.07-0.84) |

*Notes*. ABC: Activities-specific Balance Confidence scale; CI: confidence interval; FRQ: Fall Risk Questionnaire; *M<sub>Post</sub>*: mean of post measurement; *M<sub>Pre</sub>*: mean of pre measurement; SD: standard deviation; SE<sub>M diff</sub>: standard error of mean difference; SR: chair sit and reach test; STS: sit to stand test; TUG: timed up and go test. \* *p*-values reflect 1-tailed paired-samples *t*-tests (*α* = .05). The directional alternative hypotheses were *H<sub>a</sub>*: *M<sub>Post</sub>* − *M<sub>Pre</sub>* > 0 for STS, SR, and ABC (hypothesizing an increase in score) and *H<sub>a</sub>*: *M<sub>Post</sub>* − *M<sub>Pre</sub>* < 0 for TUG and FRQ (hypothesizing a reduction in score/time). Effect size directionality is standardized across measures (i.e., positive for improvement).

ABC and FRQ scores were further examined to determine the ceiling and floor effects. For the ABC, 21 participants (54%) rated their balance confidence at 90% or higher at baseline and showed a median improvement of 0.1%, whereas those scoring below 90% showed a median improvement of 6.9%. Paired-samples *t*-test of pre– and post–ABC scores in the latter group showed that this improvement was statistically significant with a large effect size (*n* = 18, *t* = – 3.30, *p* = .002, *d* = 0.78). For the FRQ, 19 participants (49%) scored within the normal range (< 4 points) at baseline and showed a median improvement of 0 points, while those scoring 4 points or above (the threshold indicating a fall risk) showed a median improvement of 1 point. Paired samples t-test of the FRQ scores in the latter group failed to reach significance (*n* = 20, *t* = 1.72, *p* = .051, *d* = 0.39).

## Discussion and Implications

The purposes of this study were to design and pilot a culturally responsive balance-enhancement exercise program for Deaf older adults. After completion of the 8-week biweekly BIM classes, older Deaf adults experienced significant improvements in physical functioning proxy measures for dynamic balance. The findings from the pilot study are encouraging.

The four foundational theories of BIM provided clear directional guidance that contributed to achieving these outcomes. The Social Ecological Model, WHO Paradigm, and TREE Inclusiveness Philosophy proved to be invaluable in the creation and delivery of a linguistically sensitive program that was responsive to Deaf cultural norms. The structural and instructional properties of BIM resonated with participants. Their engagement was evident in their attendance, on-site displays of attentiveness, and the amount and quality of debriefing feedback provided. Adherence to Exercise Training Principles also contributed to the fact that no accidents happened during the formal exercise portions of the BIM.

At the onset of the program, the majority of the participants were at high fall risk based on their vestibular impairment, lower body strength (STS), and gait speed (TUG). After completing BIM, significant changes occurred in their physical abilities. As a group, participants experienced improved functional mobility as recorded by TUG, a single performance-based metric that captures lower body strength and power, gait speed, and dynamic balance. Likewise, substantial improvements in STS were found. Standing from a seated position is an essential everyday function that requires lower body strength and stability. Additionally, study participants improved lower extremity flexibility (SR). Increased range of motion and flexibility are used in many ways, including picking objects off the floor, putting on socks and shoes, bathing or massaging feet, and gardening. In sum, it seems logical to expect gains in functional abilities tied directly to everyday activities would also bolster balance confidence.

The nearly 7% median improvement in ABC scores among participants who entered into BIM with scores below 90% is statistically significant. The minimal clinically important (MCI) difference established for ABC scores varies depending on source: 5% for older adults (American and British Geriatrics Societies’ joint Panel on Prevention of Falls in Older Persons, 2011) versus 15% for older adults seeking outpatient rehabilitation therapy (Wang et al., 2018) and 18% for adults with vestibular disorders (Wellons et al., 2022). This suggests that while on a promising trajectory, the BIM intervention may need more than eight weeks to effect meaningful change in self-perceived balance confidence.

Whilst individuals who began BIM with FRQ high-risk scores improved, the magnitude of change did not reach statistical significance. This finding may have resulted from limitations inherent in the FRQ instrument. The test includes questions beyond the control and scope of the BIM (e.g., experiencing medication side effects, reduced foot sensation, incontinence), binary response options (Yes/No) failing to capture subtle physical progress, and an item with time reference beyond BIM’s 8-week duration (“I have fallen in the past year”). Although the FRQ is a useful screening tool for identifying baseline fall risk, its content and structural constraints reduce its responsiveness as an outcome measure for short-term exercise interventions.

Even though the outcomes are encouraging, one challenge was managing unsteadiness during exercise classes and recreational activities. There were many instances where staff were able to intervene and steady participants before they fell. Nevertheless, falls occurred during recreational activities. Most falls followed vestibular-taxing quick head movements, a frequent performance demand needed to execute a chosen recreational activity. When the task requires visual focus on the fast-moving ball or shuttle, individuals with impaired vestibular function frequently experience instability regardless of their underlying physical strength and fitness. Holding onto a ping-pong table top was an effective strategy used to ameliorate instability, as was setting up the activity to be played from a seated position. Even then, participants needed to be reminded not to stand up from the seated position or shift weight on the chair too quickly (in the excitement to return a ball or birdie). In short, it became apparent that hypervigilant monitoring is warranted when managing recreational activities. There may be a benefit to grouping participants by vestibular and lower body abilities: the at-risk group may receive increased repetitive drill practices and additional balance support spotters.

To strengthen the efficacy evidence of the BIM program, future research must address several key limitations of the current study: specifically, by increasing the sample size, including a control group, and employing self-report outcome measures that are more sensitive to short-term changes. Additionally, extending the duration of both the intervention and post-program completion surveillance periods (to monitor continued engagement in exercise and balance-enhancing activities) is necessary. While it is uncertain what duration is sufficient to document balance confidence, fall risk reduction, and self-sustained exercise engagement, Lesinski et al.’s (2015) meta-analysis of 23 studies concluded exercise programs for older adults should have a dose-response of between 11-12 weeks duration. Contrastingly, Ahmed et al.’s (2024) review on intervention efficacy included 12 randomized controlled trials with durations ranging from 12 to 52 weeks.

An additional limitation of this pilot study was the requirement that participants travel to campus to participate, which introduced geographical and socio-economic sampling biases that likely influenced the results and limited generalizability. Digitally delivered, home-based fall-prevention exercise programs may be a viable pathway to overcome geographic barriers. Jasper et al.’s (2023) comparative pilot study provides preliminary evidence that functional outcomes can be achieved through teleconference meetings. Three meta-analyses of virtual reality-based exercise training or digitally delivered home-based exercises have documented effectiveness in reducing fall risk and enhancing balance, gait, mobility, and functional independence or health (Azimi Sanavi et al., 2026; Mahjur & Norasteh, 2021; Percy et al., 2023; Solis-Navarro, 2022).

Nevertheless, the benefits versus the risks of moving forward with home-based balance enhancement programs must be assessed. Participants in a digital fall prevention exercise program reported that the exercise coach was a fundamental factor in supporting their motivation and adherence (Farrell et al., 2026), suggesting the importance of effective instructors. A major concern that must be addressed before adoption of at-home exercise programs is the high rate of vestibular impairments found among Deaf older adults. In persons with complete loss of vestibular function, body movements and weight shifts while the head is in motion are destabilizing. Simply advising participants to “be careful” when exercising is inadequate. Virtual and digital exercise modalities will require careful, vigilant planning and safeguards to minimize the risk of injury while ensuring adequate aerobic and balance enhancement exercises. Additional pre-program launch initiatives include an audit of the home exercise environment (checking for uncluttered floor space, availability of counters or stable furniture to hold onto during stretching or exercise movements, especially those that require head movement), exercise footwear, and the potential need for a spotter in the room.

In summary, this pilot study demonstrates the efficacy of the BIM program in contributing to meaningful physical improvements among Deaf older adults, most notably in lower body strength, gait speed, flexibility, and dynamic balance-related mobility. A program that leads to such improvements empowers older adults to perform activities of daily living, fostering greater independence both inside and outside their homes. Moving balance interventions for older Deaf adults beyond localized pilots and into community and perhaps even digitally delivered program offerings is a necessary next step toward serving this underserved population.

## Data Availability

Data collected for this study will be available through the National Archive of Computerized Data on Aging (NACDA; openicpsr-261778).

## Acknowledgments and Data Transparency Statement

This work was supported by the National Institutes of Health – National Institute of Aging [#1 R15 AG087454-01].

The authors declare that they have no known competing financial interests or personal relationships that could have influenced the work reported in this paper.

The authors acknowledge the following individuals’ contributions to this work: Vanessa Arp (project administration [supporting], resources [supporting], supervision [supporting]); Ryan Barlongo (resources [supporting], supervision [supporting]); Tra’ Shauna Tate, Ella Gamble, Thalia Bunda, Marquize Nash, Mallory Garcia, Jacob Hartman, Olivia Armstrong, Matthew Flores, and Kate Scott (project administration [supporting], resources [supporting]); Hannah Allen, Milena Torres Bratina, Grace Kerr, Isa Yacub, Jaden Conrad, Chloe Scheff, and Alaina Milam (data collection and analysis [supporting], project administration [supporting]).

## Notes

### Competing Interest Statement

The authors have declared no competing interest.

### Clinical Trial

NCT06912607

### Author Declarations

This study was approved by Gallaudet University Institutional Review Board for Human Research (IRB-FY24-34)

## References

1. Adeyemi, O.J., Chippendale, T., Ogedegbe, O., Boatright, D., & Chodosh, J. (2025). Activity intensity and all-cause mortality following fall injury among older adults: results from a 12-year national survey. Healthcare, 13.

2. Ahmed, S., Lazo, K., McGarrigle, L., Money, A., Pendleton, N., & Todd, C. (2024). Interventions based on behavior change techniques to encourage physical activity or decrease sedentary behavior in community-dwelling adults aged 50–70: Systematic review with intervention component analysis. Journal of Aging and Physical Activity, 32(4), 554–577. 10.1123/japa.2023-0140.

3. American College of Sports Medicine. (2021). Guidelines for exercise testing and prescription. 11th ed. Wolters Kluwer.

4. Australian Sports Commission. (n.d.). TREE model. Sports Ability. https://www.ausport.gov.au/schools/resource-hub/sports_ability. Retrieved on October 30, 2026.

5. Azimi Sanavi, B., Talebpour, M., Saatchian, V., Oveysi Sani, F., & Mirakhori, F. (2026). Effects of virtual reality-based exercise training on risk of falling, walking capacity, and quality of life in healthy older adults: A systematic review and meta-analysis research. Journal of Applied Gerontology: The Official Journal of the Southern Gerontological Society, 45(7), 1223–1244. 10.1177/07334648251362512

6. Bauman, H. (2014). *DeafSpace: An architecture toward a more livable and sustainable world*. In H.-D. L. Bauman & J. J. Murray (Eds.), Deaf gain: Raising the stakes for human diversity (pp. 375–401). University of Minnesota Press.

7. Bull, F. C., Al-Ansari, S. S., Biddle, S., Borodulin, K., Buman, M. P., Cardon, G., Carty, C., Chaput, J.-P., Chastin, S., Chou, R., Dempsey, P. C., DiPietro, L., Ekelund, U., Firth, J., Friedenreich, C. M., Garcia, L., Gichu, M., Jago, R., Katzmarzyk, P. T., … Willumsen, J. F. (2020). World Health Organization 2020 guidelines on physical activity and sedentary behaviour. British Journal of Sports Medicine, 54(24), 1451–1462. 10.1136/bjsports-2020-102955

8. Centers for Disease Control and Prevention. (2017a). 30-Second Chair Stand. https://www.cdc.gov/steadi/media/pdfs/STEADI-Assessment-30Sec-508.pdf

9. Centers for Disease Control and Prevention. (2017b). Timed Up and Go (TUG). https://www.cdc.gov/steadi/media/pdfs/steadi-assessment-tug-508.pdf

10. Centers for Disease Control and Prevention. (2026, March 4). Injury center priorities. https://www.cdc.gov/injury/priorities/index.html

11. Conrad, J., Yacub, I., Milam, A., Scheff, C., Cole, J., & Tamaki, C. (2026). Validating Fall Risk Questionnaire (FRQ) in American Sign Language [Poster presentation]. American Academy of Audiology (AAA) Conference, San Antonio, TX.

12. Deandrea, S., Lucenteforte, E., Bravi, F., Foschi, R., La Vecchia, C., & Negri, E. (2010). Risk factors for falls in community-dwelling older people: a systematic review and meta-analysis. Epidemiology (Cambridge, Mass.), 21(5), 658–668. 10.1097/EDE.0b013e3181e89905

13. Deck, J., Paszkowiak, M., Clements, S., & Childress, T. (July 2026). In adults older than 65 years, does participating in the Otago exercise program decrease fear of falling versus no program participation? Evidence-Based Practice 29(7), 16–17.

14. Diamond, L., Izquierdo, K., Canfield, D., Matsoukas, K., & Gany, F. (2019). A systematic review of the impact of patient-physician non-English language concordance on quality of care and outcomes. Journal of General Internal Medicine, 34(8), 1591–1606. 10.1007/s11606-019-04847-5

15. Dolan, H., Pohl, J., Pituch, K., & Coon, D. (2025). Perceived balance predicts falls in community-dwelling older adults. Innovation in Aging, 8(Suppl 1), 840. 10.1093/geroni/igae098.2720

16. Farrell, S., Bajdek, N., Dishaw, M., Garabedian, P., Williams, A., Hachen, N., Valderrábano, R., Reid, K., & Latham, N. (2026). A Qualitative Analysis of a Digital Fall Prevention Exercise Program for Older Adults With Increased Fall Risk. Journal of Applied Gerontology, 45(2), 393–403. 10.1177/07334648251342833

17. Faul, F., Erdfelder, E., Buchner, A., & Lang, A.-G. (2009). Statistical power analyses using G*Power 3.1: Tests for correlation and regression analyses. Behavior Research Methods, 41(4), 1149–1160. 10.3758/BRM.41.4.1149

18. Faul, F., Erdfelder, E., Lang, A.-G., & Buchner, A. (2007). G*Power 3: A flexible statistical power analysis program for the social, behavioral, and biomedical sciences. Behavior Research Methods, 39(2), 175–191. 10.3758/BF03193146

19. Freiberger, E. (2022). Falls prevention: Adherence, fear of falling, assessment, and intervention. Journal of Aging and Physical Activity, 30(4), 569–571. 10.1123/japa.2022-0101

20. Garnett, M. F., Weeks, J. D., & Zehner, A. M. (2025). Unintentional fall deaths in adults age 65 and older: United States, 2023. National Center for Health Statistics. 10.15620/cdc/174601

21. Harvard Health Publishing. (2015). Walking for health.

22. Harvard Health Publishing. (2019). Strength and power training for older adults: two complete workouts to start rebuilding your muscles.

23. Harvard Health Publishing. (2022). Better balance: Simple exercises to improve stability and prevent falls.

24. Harvard Health Publishing. (2023). Stretching: Stretches for the whole body to improve flexibility and reduce pain.

25. Harvard Health Publishing. (2024a). Discovering functional fitness: Exercise to keep you active as you age, make everyday tasks easier, and even improve your game.

26. Harvard Health Publishing. (2024b). Everyday balance exercises: Easy moves to get stronger, feel steadier, and stay active.

27. Harvard Health Publishing. (2024c). Preventing falls: Strategies to maintain health, strength, balance, and a safe home environment.

28. Hernán-Santana, G., & Sobrido-Prieto, M. (2026). Effectiveness of group exercise on the quality of life in older adults: A systematic review. Journal of Geriatric Physical Therapy 10.1519/JPT.0000000000000502

29. The Jamovi Project (2025). *Jamovi* (Version 2.7). [Computer software]. https://jamovi.org

30. Jasper, A., Chamberlain, D., & Lazaro, R. (2023). Teleassessment of balance and gait outcome measures in older adults: A systematic review. Topics in Geriatric Rehabilitation, 39(4), 288–293. 10.1097/TGR.0000000000000412

31. Jones, C. A., Siever, J., Knuff, K., Van Bergen, C., Kurtz, D., & Miller, H. (2019). Walk, Talk and Listen: A pilot randomised controlled trial targeting functional fitness and loneliness in older adults with hearing loss. BMJ Open, 9(8), e026169. 10.1136/bmjopen-2018-026169

32. Koon, L. M., Remillard, E. T., & Rogers, W. A. (2025). Understanding everyday challenges of aging with disability through lived experiences. The Gerontologist, 65 (7), gnaf129. doi10.1093/geront/gnaf129

33. Krawczyk, T., Piasecki, J., Wasylewski, M., & Waligora, M. (2024). Ethics of research engagement with Deaf people. A qualitative evidence synthesis. Journal Of Deaf Studies And Deaf Education, 29(4), 443–455. 10.1093/jdsade/enae024

34. Lambert, J., Ghadry-Tavi, R., Knuff, K., Jutras, M., Siever, J., Mick, P., Roque, C., Jones, G., Little, J., Miller, H., Van Bergen, C., Kurtz, D., Murphy, M. A., & Jones, C. A. (2017). Targeting functional fitness, hearing and health-related quality of life in older adults with hearing loss: Walk, Talk ‘n’ Listen, study protocol for a pilot randomized controlled trial. Trials, 18(1), Article 47. 10.1186/s13063-017-1792-z

35. Lesinski, M., Hortobágyi, T., Muehlbauer, T., Gollhofer, A., & Granacher, U. (2015). Effects of balance training on balance performance in healthy older adults: A systematic review and meta-analysis. *Sports Medicine (Auckland*, N.Z.), 45(12), 1721–1738. 10.1007/s40279-015-0375-y

36. Lin, F. R., & Ferrucci, L. (2012). Hearing loss and falls among older adults in the United States. Archives of Internal Medicine, 172(4), 369–371. 10.1001/archinternmed.2011.728

37. Liu, C.-J., Chang, W.-P., Shin, Y. C., & Su, C. H. (2024). Is functional training functional? A systematic review of its effects in community-dwelling older adults. European Review of Aging and Physical Activity, 21(1), Article 32. 10.1186/s11556-024-00366-3

38. Mahjur, M., & Norasteh, A. A. (2021). The effect of unsupervised home-based exercise training on physical functioning outcomes in older adults: A systematic review and meta-analysis of randomized controlled trials. Biological research for nursing, 23(3), 504–512. 10.1177/1099800421989439

39. McKee, M., Thew, D., Starr, M., Kushalnagar, P., Reid, J. T., Graybill, P., Velasquez, J., & Pearson, T. (2012). Engaging the Deaf American Sign Language community: Lessons from a community-based participatory research center. *Progress in Community Health Partnerships: Research*, Education, and Action, 6(3), 321–329. 10.1353/cpr.2012.0037

40. McLeroy, K. R., Bibeau, D., Steckler, A., & Glanz, K. (1988). An ecological perspective on health promotion programs. Health Education Quarterly, 15(4), 351–377. 10.1177/109019818801500401

41. Montero-Odasso, M., van der Velde, N., Martin, F. C., Petrovic, M., Tan, M. P., Ryg, J., Aguilar-Navarro, S., Alexander, N. B., Becker, C., Blain, H., Bourke, R., Cameron, I. D., Camicioli, R., Clemson, L., Close, J., Delbaere, K., Duan, L., Duque, G., Dyer, S. M., Freiberger, E., … Task Force on Global Guidelines for Falls in Older Adults (2022). World guidelines for falls prevention and management for older adults: A global initiative. Age and Ageing, 51(9), afac205. 10.1093/ageing/afac205

42. Panel on Prevention of Falls in Older Persons, American Geriatrics Society and British Geriatrics Society (2011). Summary of the Updated American Geriatrics Society/British Geriatrics Society clinical practice guideline for prevention of falls in older persons. Journal of the American Geriatrics Society, 59(1), 148–157. 10.1111/j.1532-5415.2010.03234.x

43. Peng, R., Chang, J., Zhu, Y., Yang, R., Feng, H., & Zhao, Y. (2026). Exercise habits in older adults: A scoping review. *Healthcare (Basel*, Switzerland), 14(15), 2375. 10.3390/healthcare14152375

44. Percy, D., Phillips, T., Torres, F., Chaleunphonh, M., & Sung, P. (2023). Effectiveness of virtual reality-based balance and gait in older adults with fear of movement: A systematic review and meta-analysis. Physiotherapy Research International, 28(4), e2037. 10.1002/pri.2037

45. Pillay, J., Gaudet, L. A., Saba, S., Vandermeer, B., Ashiq, A. R., Wingert, A., & Hartling, L. (2024). Falls prevention interventions for community-dwelling older adults: systematic review and meta-analysis of benefits, harms, and patient values and preferences. Systematic reviews, 13(1), 289. 10.1186/s13643-024-02681-3

46. R Core Team (2025). R: A language and environment for statistical computing. (Version 4.5) [Computer software]. Retrieved from https://cran.r-project.org. (R packages retrieved from CRAN snapshot 2025-05-25).

47. Rikli, R. E., & Jones, C. J. (2013). Senior Fitness Test Manual (2nd ed.). Human Kinetics.

48. Sadaqa, M., Németh, Z., Makai, A., Prémusz, V., & Hock, M. (2023). Effectiveness of exercise interventions on fall prevention in ambulatory community-dwelling older adults: A systematic review with narrative synthesis. Frontiers in Public Health, 11, 1209319. 10.3389/fpubh.2023.1209319

49. Sallis, J. F., Owen, N., & Fisher, E. B. (2015). Ecological models of health behavior. In K. Glanz, B. K. Rimer, & K. Viswanath (Eds.), Health Behavior: Theory, Research, and Practice (5th ed., pp. 43–64). Jossey-Bass.

50. Solis-Navarro, L., Gismero, A., Fernández-Jané, C., Torres-Castro, R., Solá-Madurell, M., Bergé, C., Pérez, L. M., Ars, J., Martín-Borràs, C., Vilaró, J., & Sitjà-Rabert, M. (2022). Effectiveness of home-based exercise delivered by digital health in older adults: a systematic review and meta-analysis. Age and Ageing, 51(11), afac243. 10.1093/ageing/afac243

51. Tamaki, C. & Cole, J. (2026). Activities-Specific Balance Confidence Scale (ABC) in American Sign Language. Journal of the American Academy of Audiology, 00:1–10. 10.3766/jaaa.250115

52. U.S. Department of Health and Human Services, Office of Disease Prevention and Health Promotion. (n.d.). Reduce the rate of emergency department visits due to falls among older adults—OA-03. Healthy People 2030. https://odphp.health.gov/healthypeople/objectives-and-data/browse-objectives/older-adults/reduce-rate-emergency-department-visits-due-falls-among-older-adults-oa-03

53. U.S. Department of Health and Human Services. (2018). Physical activity guidelines for Americans (2nd ed.). https://health.gov/sites/default/files/2019-09/Physical_Activity_Guidelines_2nd_edition.pdf

54. Wang, Y. C., Sindhu, B., Lehman, L., Li, X., Yen, S. C., & Kapellusch, J. (2018). Rasch analysis of the Activities-Specific Balance Confidence Scale in older adults seeking outpatient rehabilitation services. The Journal of Orthopedic & Sports Physical Therapy, 48(7), 574–583. 10.2519/jospt.2018.8023

55. Wellons, R. D., Duhe, S. E., MacDowell, S. G., Hodge, A., Oxborough, S., & Levitzky, E. E. (2022). Estimating the minimal clinically important difference for balance and gait outcome measures in individuals with vestibular disorders. Journal of Vestibular Research: Equilibrium & Orientation, 32(3), 223–233. 10.3233/VES-201630

56. Wollesen, B., Pocovi, N. C., Salvestro, K., Hurley, S., Seydell, L., Scrivener, K., & Dean, C. M. (2021). Multitask training to improve walking performance in older adults with hearing impairment: A feasibility study. Aging and Health Research, 1(3), 100028. 10.1016/j.ahr.2021.100028

57. World Health Organization. (2020). Decade Of Healthy Ageing: Plan Of Action 2020–2030. https://cdn.who.int/media/docs/default-source/decade-of-healthy-ageing/decade-proposal-final-apr2020-en.pdf

58. World Health Organization. (2021, April 26). *Falls*. https://www.who.int/news-room/fact-sheets/detail/falls

59. World Health Organization. (2023). Progress report on the United Nations Decade of Healthy Ageing, 2021–2023.

60. Yeo, B. S. Y., Tan, V. Y. J., Ng, J. H., Tang, J. Z., Sim, B. L. H., Tay, Y. L., Chowdhury, A. R., David, A. P., Jiam, N. T., Kozin, E. D., & Rauch, S. D. (2025). Hearing loss and falls: A systematic review and meta-analysis. JAMA Otolaryngology–Head & Neck Surgery, 151(5), 485–494.

